# Oil Immersed Lossless Total Analysis System (OIL-TAS): Integrated RNA Extraction and Detection for SARS-CoV-2 Testing

**DOI:** 10.1101/2020.09.30.20204842

**Authors:** Duane S. Juang, Terry D. Juang, Dawn M. Dudley, Christina M. Newman, Thomas C. Friedrich, David H. O’Connor, David J. Beebe

**Author notes:** These authors contributed equally to this work (Duane S. Juang, Terry D. Juang).

## Abstract

The coronavirus disease 2019 (COVID-19) pandemic exposed difficulties in scaling current quantitative PCR (qPCR)-based diagnostic methodologies for large-scale infectious disease testing. Bottlenecks include the lengthy multi-step process of nucleic acid extraction followed by qPCR readouts, which require costly instrumentation and infrastructure, as well as reagent and plastic consumable shortages stemming from supply chain constraints. Here we report a novel Oil Immersed Lossless Total Analysis System (OIL-TAS), which integrates RNA extraction and detection onto a single device that is simple, rapid, cost effective, uses minimal supplies and requires reduced infrastructure to perform. We validated the performance of OIL-TAS using contrived samples containing inactivated SARS-CoV-2 viral particles, which show that the assay can reliably detect an input concentration of 10 copies/μL and sporadically detect down to 1 copy/μL. The OIL-TAS method can serve as a faster, cheaper, and easier-to-deploy alternative to current qPCR-based methods for infectious disease testing.

## Main

Broad testing is crucial for monitoring and controlling the spread of infectious disease outbreaks. The COVID-19 pandemic caused by severe acute respiratory syndrome coronavirus 2 (SARS-CoV-2) spread rapidly around the world after its initial outbreak from Wuhan China in December 2019^1,2^, quickly overwhelming current diagnostic testing capacity and supply chains. The go-to gold standard diagnostic technique for novel infectious diseases is usually molecular-based (i.e. quantitative polymerase chain reaction (qPCR)), owing to the relative ease to develop a highly sensitive and specific test within a short timeframe. However, existing methodologies for molecular testing have proven difficult to scale, owing to the assay’s complexity, lengthy operation, requirement of dedicated centralized testing infrastructure, and supply chain constraints.

The current standard method for SARS-CoV-2 nucleic acid testing is a multi-step protocol involving RNA extraction from patient samples (mostly commonly nasal, nasopharyngeal or oropharyngeal swabs, and saliva) using magnetic bead-based or column-based methods, followed by reverse transcription quantitative PCR (RT-qPCR) based detection of the extracted RNA. The RNA extraction process typically involves: 1) mixing the sample with lysis/binding buffer, 2) solid-phase capture of the viral RNA via magnetic beads or columns, 3) multiple washes involving magnetic separation or centrifugation for each wash, 4) elution of viral RNA with water or a low salt buffer, 5) adding the eluted RNA to a PCR plate containing RT-qPCR master mix and primers, followed by thermocycling and data capture in a qPCR machine. This process often takes up to 4 hours and is challenging to scale because of the complexity of the RNA extraction process, coupled with the RT-qPCR process itself which requires over 1 hour of on-machine real-time fluorescence measurements, significantly limiting assay turnaround time. The nature of this complex multi-step process also necessitates the use of a significant amount of plastic consumables (pipet tips, tubes, plates, columns, etc.) which become biohazardous waste after the assay. For example, it is estimated that up to 10 pipet tips (6 for RNA extraction, 4 for PCR setup) are consumed per sample for a SARS-CoV-2 qPCR-based molecular test^3^. This has also led to widespread shortages of essential plastic consumables to perform the tests, significantly bottlenecking testing capacity^4^.

To address these challenges for scaling up testing, here we report an alternative molecular assay we termed the Oil Immersed Lossless Total Analysis System (OIL-TAS) which integrates RNA extraction and detection into a single device with the footprint of a generic 96-well plate. The OIL-TAS has advantages over RT-qPCR including simplicity, lossless sample handling, less reagent/plastic consumable consumption, lower cost, good sensitivity, and increased speed and throughput. Importantly, the OIL-TAS can be operated using pipettes, a shaker, an oven, and an image capture device, which are widely available in biomedical laboratories without the need for costly or specialized instruments. We also deliberately engineered the assay to be compatible with open source, non-proprietary reagents and employ a colorimetric isothermal detection method to reduce assay time and avoid adding burden to current qPCR testing supply chains and clinical workflows.

The OIL-TAS assay integrates three technologies: 1) an underoil droplet microfluidic technology called Exclusive Liquid Repellency (ELR) that allows for lossless sample processing^5,6^; 2) a rapid solid-phase analyte extraction method called Exclusion-based Sample Preparation (ESP)^7–12^; and 3) isothermal amplification with colorimetric readout (Loop-Mediated Isothermal Amplification, LAMP^13,14^) resulting in a simple, high throughput, low cost test that could be quickly and broadly implemented using simple tools and consumables that are widely available. The OIL-TAS device builds upon our previously reported ELR technology, which describes physical conditions where an aqueous droplet can be fully repelled from a solid surface (contact angle = 180°) in the presence of an oil phase when a specific set of oil and solid interfacial energy properties are met. The conditions in which ELR will occur are when the sum of the interfacial energies of the solid/oil and aqueous/oil interface are equal to, or less than the solid/aqueous interfacial energy. Through experimentation, we found that this condition can be accomplished by employing a Polydimethylsiloxane (PDMS)-silane functionalized surface paired with silicone oil as the oil phase^15^. The most significant advantage of ELR is that it prevents adsorption of biological samples to surfaces and thus has very little, if any, associated sample loss. With ELR, one can create individually isolated aqueous droplets immersed under a common oil phase, each droplet providing a completely isolated reaction condition with no crosstalk with the solid surface of the reservoir, enabling liquid handling without loss.

Although ELR can effectively mitigate surface adsorption-mediated sample loss, another common source of loss during sample processing often occurs during solid-phase extraction processes: namely, target analytes (such as RNA) that are bound to a solid phase (magnetic beads or column resin) can fall off prematurely during the multiple washing steps that are common in solid-phase extraction processes such as RNA extraction. This issue can be effectively reduced by employing Exclusion-based Sample Preparation (ESP), which refers to a collection of solid-phase analyte extraction techniques by which analytes bound to a solid-phase (magnetic beads) can be extracted out of a complex sample by transporting the beads via a magnet through an immiscible interface (oil or air) to “exclude” non-target contaminants from the sample. The ESP process replaces the multiple washing operations in traditional solid-phase extraction techniques with a simple magnetic dragging operation through the immiscible interfaces, resulting in a much shorter processing time and higher sample recovery^16^.

OIL-TAS combines the ELR and ESP technologies into a rapid and flexible integrated analyte extraction and detection platform. The OIL-TAS consists of an array of immobilized ELR droplets immersed under an oil bath, where each droplet can contain a sample, wash solution, or reaction solution (Fig. 1). The oil phase provides droplet isolation and liquid repellent properties, and also serves as a water-immiscible extraction interface for performing ESP: magnetic beads can be added to the aqueous droplets and “extracted” from one droplet to another by dragging a magnet across the bottom of the device for analyte purification (Fig. 1 and Supplementary Video 1). Additionally, the oil overlay also provides many other advantages including: 1) It prevents evaporation of droplets, thus enabling reactions that require heating (such as isothermal amplification) to be performed using much smaller volumes, 2) It prevents cross-contamination of reagents/samples and LAMP-amplified products, 3) It prevents aerosol formation during operation, as aerosols would be effectively trapped under the oil overlay, 4) It prevents contamination from the environment, as dust particles/other contaminants would be shielded by the oil overlay, and 5) It enables longer-term storage of individual aqueous reagents in the plate via freezer storage (−20 °C).

**Fig. 1.**
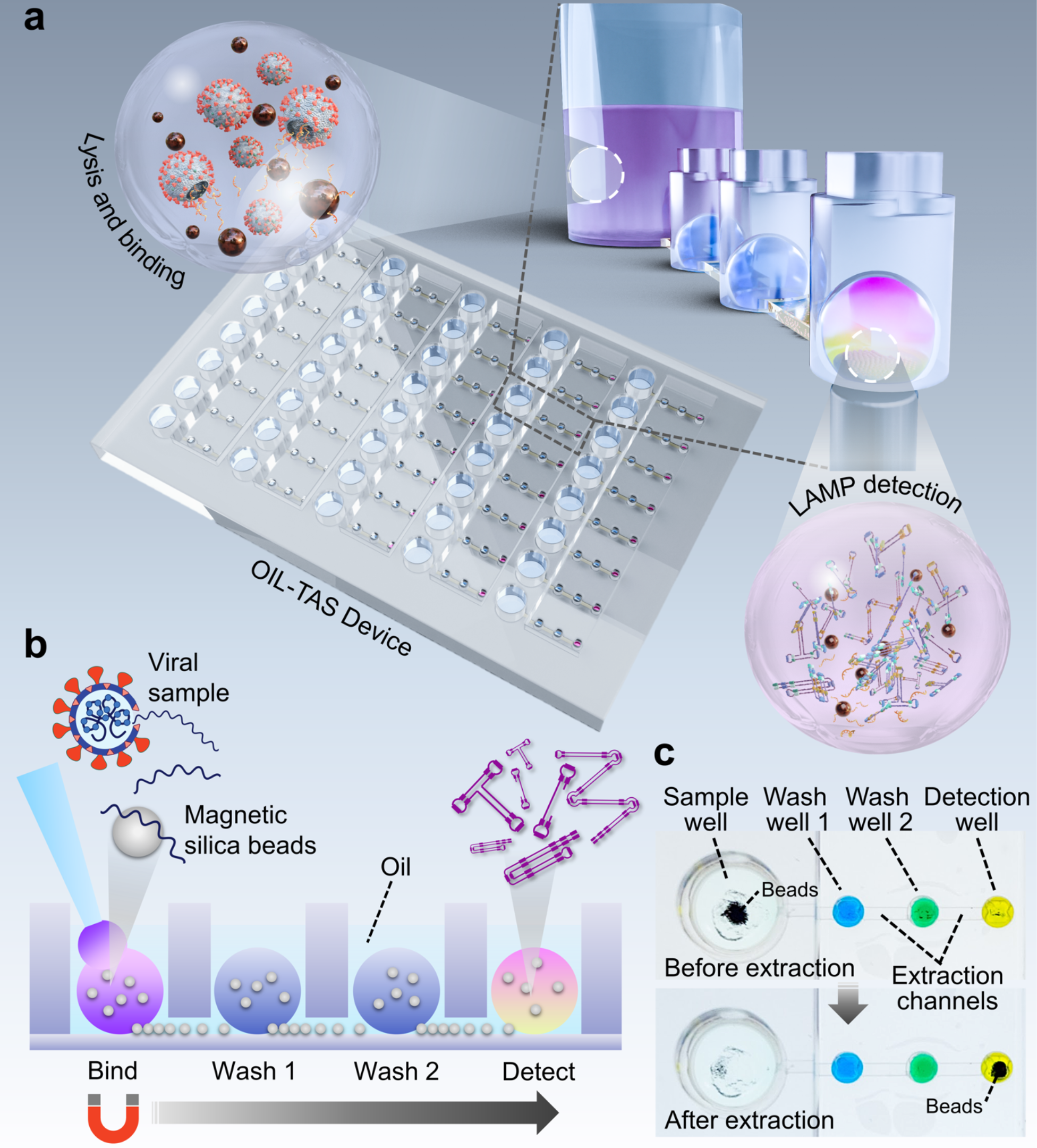
Integrated RNA extraction and detection with the OIL-TAS platform. **a** Schematic depicting the design and essential components of the OIL-TAS. **b** Side view cartoon illustrating the operation principle of OIL-TAS sample extraction. Owing to the exclusive liquid repellent (ELR) nature of the device surface, droplets appear spherical which minimizes contact with the device surface and hence biofouling associated sample loss. **c** Images of a single unit of the OIL-TAS device before and after extraction. Each unit includes a large sample well for sample lysis and binding, followed by two washing wells and a detection well. Interconnecting the wells are oil-filled extraction channels. Wells are filled with droplets of food coloring for visualization.

In order to robustly immobilize the spherical ELR droplets, we designed an array of wells for trapping the droplets, with shallow extraction channels interconnecting the wells for performing extraction (Fig. 1). One extraction unit of the device consists of a large sample well connected to 3 small wells (for wash 1, wash 2, and detection) (Fig. 1b and 1c). Each device the size of a microtiter plate (127.76 mm x 85.48 mm) contains 40 extraction units which allows for the simultaneous parallel processing of 40 samples. The whole device is treated using ELR chemistry, which ensures that any given surface of the device is fully repellent to aqueous media to prevent biomolecule adsorption. Adding silicone oil into the device wells fills the wells and extraction channels simultaneously with oil via capillary action. When aqueous droplets are pipetted into the oil-filled wells, it forms an array of oil-immersed aqueous droplets separated by oil-filled extraction channels (Fig. 1). The device is fabricated from 3 sheets of heat-resistant, optically transparent plastic (polycarbonate, Lexan) which provides advantages including a wide working temperature range (−40 °C to 115 °C), uniform chemical properties throughout the device, and compatibility with mass manufacturing injection molding processes (Fig. 2a). The optically clear thin device bottom (127 μm) allows for efficient magnetic manipulation, rapid heat transfer, and optical access, making the device compatible with visual, absorbance, fluorescence, luminescence, and microscopy-based readouts. The vertical pitch of the wells is 9 mm to enable parallel operation via a standard multichannel pipet, whereas the horizontal pitch is 4.5 mm (corresponding to a conventional 384-well plate) to enable data acquisition via microplate readers. The inlets of the small wells (wash well 1, wash well 2, and detection well) incorporate a non-circular collar to help align the pipet tip during liquid dispensing (Fig. 2b). The non-circular collar design can prevent the circular pipet tip from forming a tight seal over the inlet, allowing oil to escape over the top of the well when being displaced by pipetting aqueous media into the well. The collar also prevents the aqueous droplet from accidental escape from the top of the well, as long as the diameter of the dispensed droplet is larger than the opening of the collar.

**Fig. 2.**
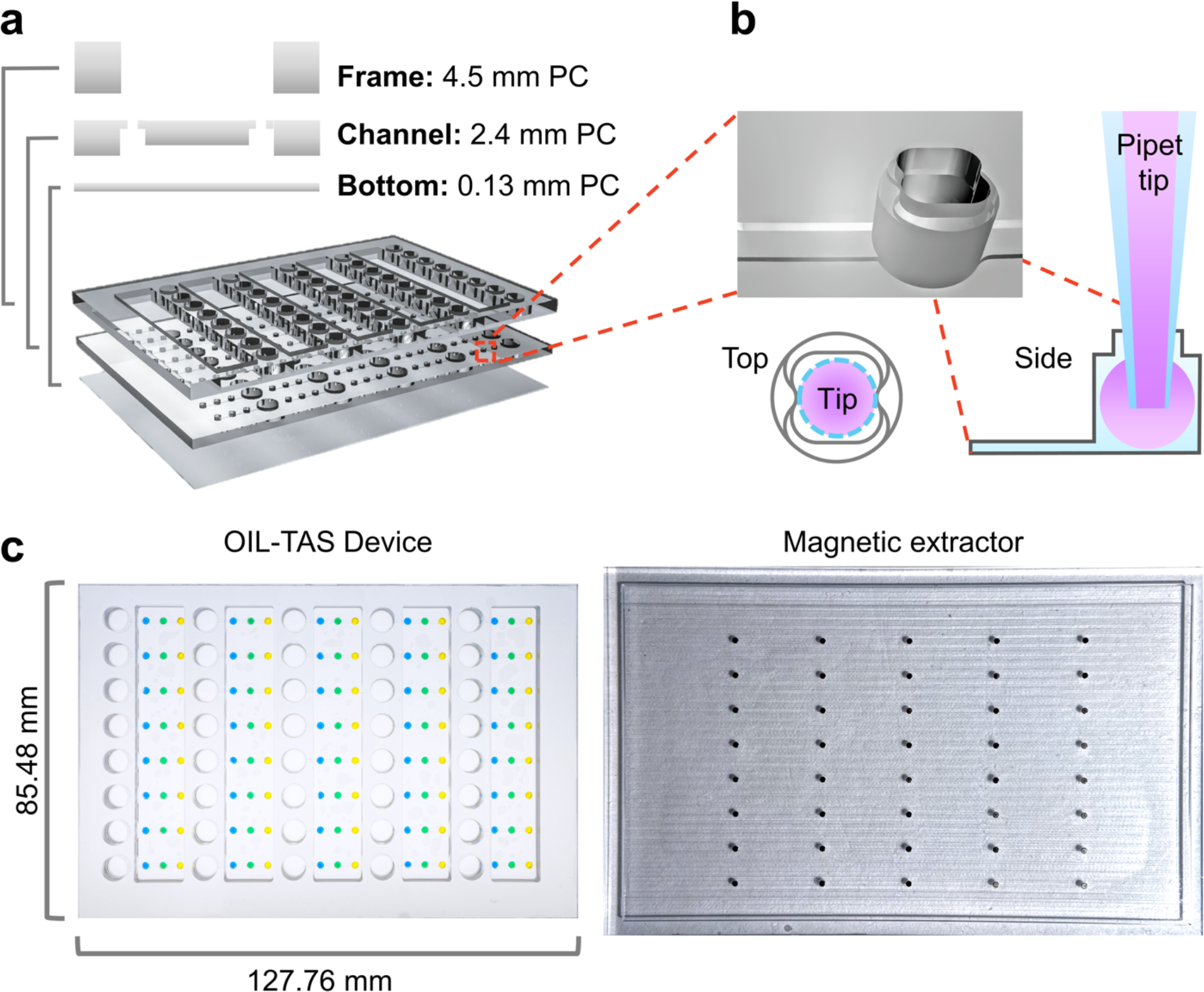
Design and assembly of the OIL-TAS device. **a** Exploded view of the OIL-TAS device consisting of 3 layers fabricated from polycarbonate (PC). **b** Design of well collar for pipet tip alignment and preventing droplet escape. **c** Images of OIL-TAS device (left) and magnetic extractor (right). Food coloring was added to the wells to facilitate visualization.

When using the OIL-TAS device for SARS-CoV-2 testing, the number of operation steps is minimized to 1) add oil, washing solutions, and LAMP reaction solution into the device; 2) mix sample and lysis buffer/beads in a plate; 3) transfer the bead/sample solution to the device via a pipet, and perform extraction by dragging a magnet across the bottom of the device, 4) place the device in an oven for isothermal amplification, and 5) data readout via visual inspection or an image acquisition device (flatbed scanner, camera/smartphone, or plate reader) (Supplementary Fig. 1). An operator can perform the whole process manually in less than 30 min for a 40-sample device (excluding the isothermal amplification time), although this can be significantly shortened by using reagent pre-loaded plates and an automated liquid handling system. This compares favorably to commercial automated RNA extraction systems, which commonly take between 45∼120 min to process 96 samples^17–19^.

To quantify the sample purification performance of the OIL-TAS method, we evaluated the amount of liquid carryover from one droplet to another during the magnetic extraction process in the OIL-TAS device using a fluorescent molecule (acridine orange) as a model contaminant (Supplementary Fig. 2). Results show only 46.4 nL of aqueous solution was carried over from one droplet to another during the magnetic extraction process (equivalent to 1.16% of the 4 μL droplet), which means that approx. ∼0.00015% of contaminating solution will remain after the 3 sequential extractions (Supplementary Fig. 2). We also tested a variety of channel heights ranging between 100 μm to 600 μm and found that magnetic extraction works throughout this broad range of channel heights (Supplementary Video 2). However, to prevent bead clogging during extraction or accidental droplet escape through the extraction channels during device handling, we selected an optimized height of 200 μm and a width of 800 μm for the extraction channels. We validated that the droplets in this design were highly stable without any observed escape or cross contamination after rocking on a rocking platform shaker at 30 rpm for 30 min (Supplementary Fig. 3 and Supplementary Video 3) or even after vigorous shaking on an orbital plate shaker at 900 rpm (Supplementary Video 3).

To avoid the shortage and supply chain issues with scaling up testing for SARS-CoV-2, we designed our assay to 1) be compatible with completely open source, non-proprietary materials and reagents for RNA extraction, and 2) use very little (4 μL) proprietary reagent (colorimetric LAMP master mix) per reaction to conserve on limited materials and reduce cost. A main bottleneck in RNA extraction supplies is the proprietary nature of most reagents in commercial RNA extraction kits, including the lysis/binding buffer, washing buffers, as well as magnetic beads, which constrains supply to the production capacity of a given company. However, it is known that the majority of commercial solid-phase RNA extraction methods employ variants of a guanidine salt/silica binding chemistry reported in 1990 by Boom et al^20^. RNA binds to silica surfaces (such as silica columns or magnetic silica beads) under high guanidine salt conditions in the presence of an alcohol (commonly ethanol or isopropanol), which are the major active ingredients in RNA lysis/binding buffers. There have been efforts to develop various open source RNA lysis/binding buffer recipes from the scientific community^21–25^, which were demonstrated by the authors to be similar in performance to commercial proprietary kits. We adopted a lysis buffer recipe reported by Escobar et al.^21^ with slight modifications (recipe in Methods section) and found it to perform well in our system. We also tested magnetic silica beads from different vendors (MagAttract beads from Qiagen, MagneSil beads from Promega, MagBinding beads from Zymo Research, and SeraSil-Mag beads from Cytiva) for compatibility with this open source RNA extraction chemistry and our OIL-TAS system. We found that the beads from these vendors were all compatible with OIL-TAS extraction process and successfully extracted RNA from gamma-irradiated inactivated SARS-CoV-2 viral particles spiked into viral transport media (VTM) to yield good sensitivity via LAMP detection with an N gene primer set^26^ targeting SARS-CoV-2 (Fig. 3). We selected the N Gene primer set reported by Broughton et al.^26^ because it was granted emergency use authorization (EUA) by the U.S. Food and Drug Administration for Mammoth Biosciences^27^ and Color Genomics^28^ for SARS-CoV-2 testing. We did observe minor differences in magnetic responsiveness and performance between the beads from different vendors, although minor optimizations for each bead type may further improve performance. Qiagen MagAttract beads were selected for following experiments due to their higher magnetic responsiveness and sensitivity.

**Fig. 3.**
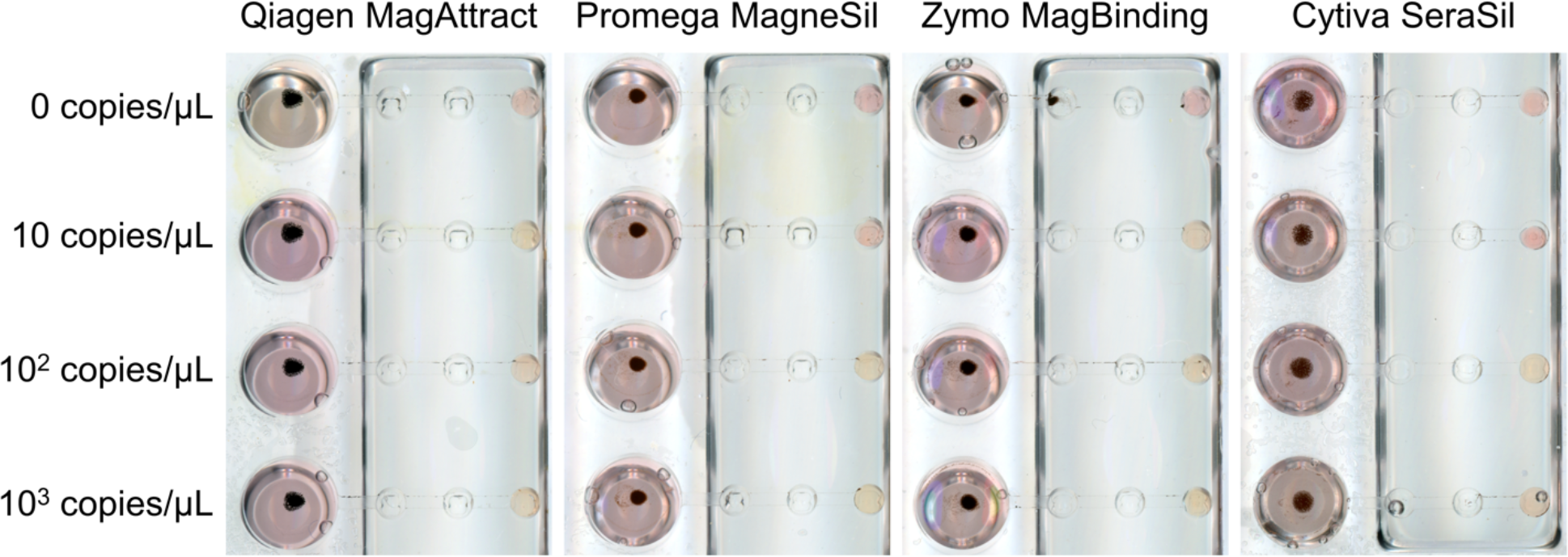
Integrated extraction and LAMP detection of γ-irradiated inactivated SARS-CoV-2 particles (strain USA-WA1/2020) from the Biodefense and Emerging Infections Research Resources Repository (BEI) with OIL-TAS. The OIL-TAS method is compatible with open source RNA extraction buffers and magnetic silica beads from various vendors for integrated RNA extraction and LAMP detection of SARS-CoV-2.

To further improve on-site assay throughput and operation simplicity, we also tried preloading the OIL-TAS device to contain most of the necessary assay reagents (oil, washing solutions, and LAMP master mix) followed by freezer storage (−20 °C). We found that after a week-long storage at −20 °C (longer periods were not tested), the OIL-TAS device still retained similar performance after the freeze-thaw compared to a freshly prepared device (Supplementary Fig. 4), suggesting the potential for pre-packaging assays into a ready-to-run format in OIL-TAS.

We further validated the performance and sensitivity of the assay using replication deficient viral particle samples (SeraCare AccuPlex SARS-CoV-2 Verification Panel) in viral transport media. Results show that we were able to detect down to 1 copy/μL (Fig. 4a, left panel) using the Broughton et al. N Gene primer set. The assay also showed no false positives using the RNase P negative extraction control samples included in the SeraCare AccuPlex kit. We observed similar sensitivity of the assay using BEI gamma-irradiated inactivated SARS-CoV-2 viral particles spiked into VTM (Fig. 4a, right panel). The analytical limit of detection (LoD) of the assay for SARS-CoV-2 viral particles was evaluated by 3 serial 10-fold dilutions of gamma-irradiated viral particles with 10 replicates. Results show that we were able to detect 6/10 samples for 1 copy/μL, and 10/10 for 10 copies/μL and 100 copies/μL concentrations (Fig. 4b and Fig. 4c), with none detected (0/10) for 0 copies/μL. These results show that our assay can reliably detect down to 10 copies/μL and approach 1 copy/μL sensitivity, with good specificity.

**Fig. 4.**
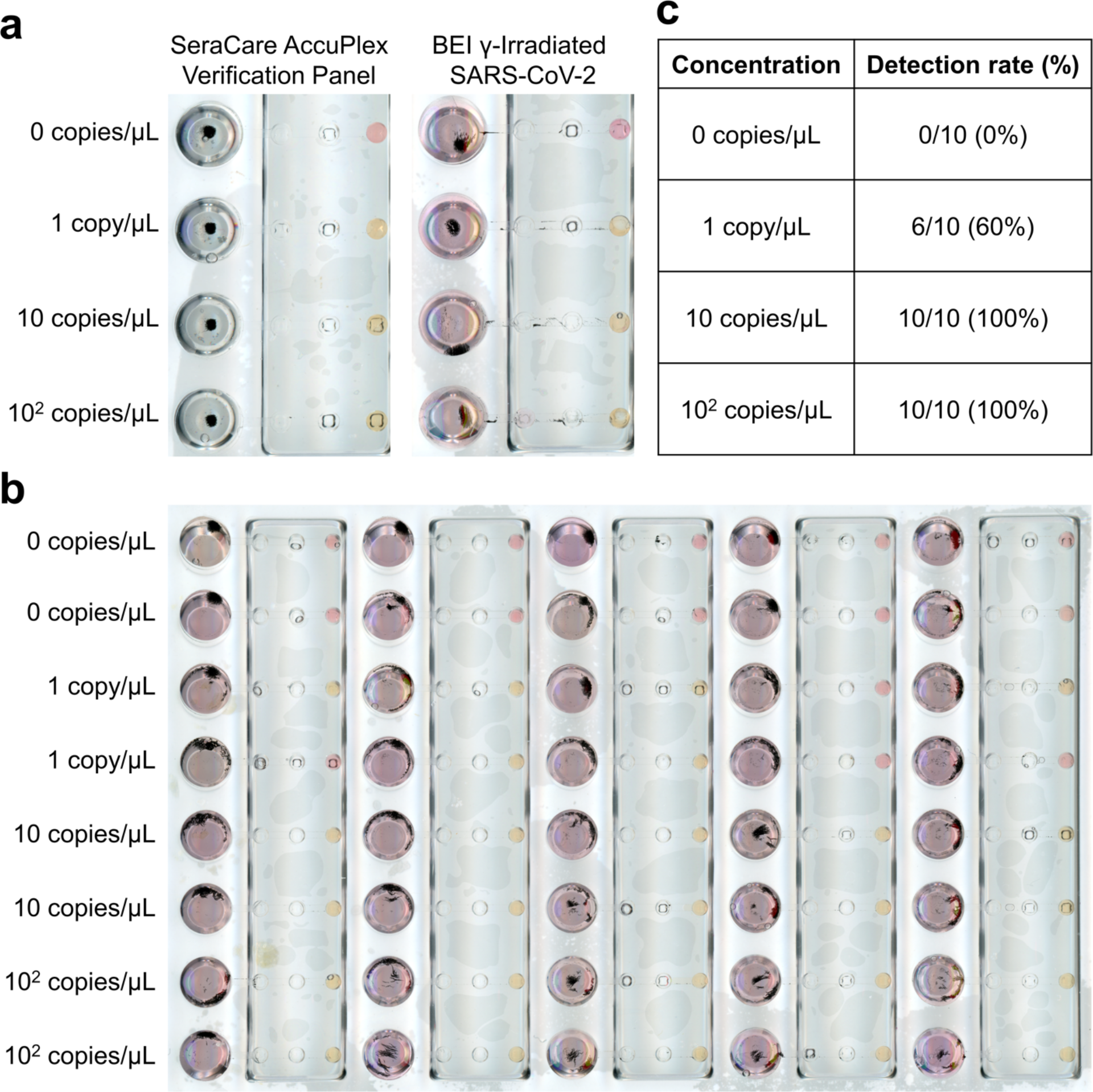
Extraction and detection of contrived SARS-CoV-2 viral particle samples using OIL-TAS. **a** RNA extraction and detection of viral particles. Left, synthetic viral particles containing SARS-CoV-2 consensus sequences (AccuPlex verification panel, SeraCare Life Sciences Inc.). Right, γ-irradiated inactivated SARS-CoV-2 particles (strain USA-WA1/2020) from BEI. **b** Sensitivity of the OIL-TAS technique for detecting SARS-CoV-2 viral particles across 10 replicates. **c** Detection rate of the OIL-TAS technique across different viral particle concentrations (quantified from panel **b**).

Most nucleic acid-based diagnostic assays include an extraction (process) control readout in addition to the target sequence to validate that the assay is performing correctly. Thus, we designed a single input, dual output version of the OIL-TAS device (Fig. 5a) to enable simultaneous multiplexed readout from a single sample to increase the robustness of the assay. The dual output device footprint remains the same and also has the same number of input sample wells (40 samples) as the single output device, but the number of small wells (wash wells and detection wells) were doubled (Fig. 5b). This was accomplished by halving the horizontal pitch between the small wells to 4.5 mm, corresponding to that of a 384-well plate, and increasing the size of the sample wells to accommodate larger sample volumes. We employed this design for the simultaneous detection of the N gene from SARS-CoV-2 and human RNase P as an extraction process control, or the simultaneous detection of two genomic targets from SARS-CoV-2 (N gene and ORF1a). This was done by using different primer sets in the 2 output wells, including primers for the N gene targeting SARS-CoV-2^26^, As1e targeting the open reading frame (Orf1a) of SARS-CoV-2^29^, and RNase P targeting human cells as a process control^30^. We spiked human A549 cells at a concentration of 10 cells/μL into VTM containing 0, 1, 10, and 100 copies/μL inactivated SARS-CoV-2 particles as a contrived sample. Results show that the sensitivity of the assay was similar with the dual output device, detecting down to 1 copy/μL SARS-CoV-2 viral particles, with all the RNase P extraction controls positive (Fig. 5c). Multiplexed detection of SARS-CoV-2 using two primers also show similar sensitivity (Fig. 5d). Worth mentioning is that at 1 copy/μL concentrations, our detection rate was 6/10 using the single output device (Fig. 4c), but the sensitivity and reliability can be potentially improved in the dual output device as 1 of the 2 primer sets can still detect a positive signal (Fig. 5d). Worth noting is that the number of output wells (and hence multiplexing targets) may be scaled up with just minor modifications to the current design.

**Fig. 5.**
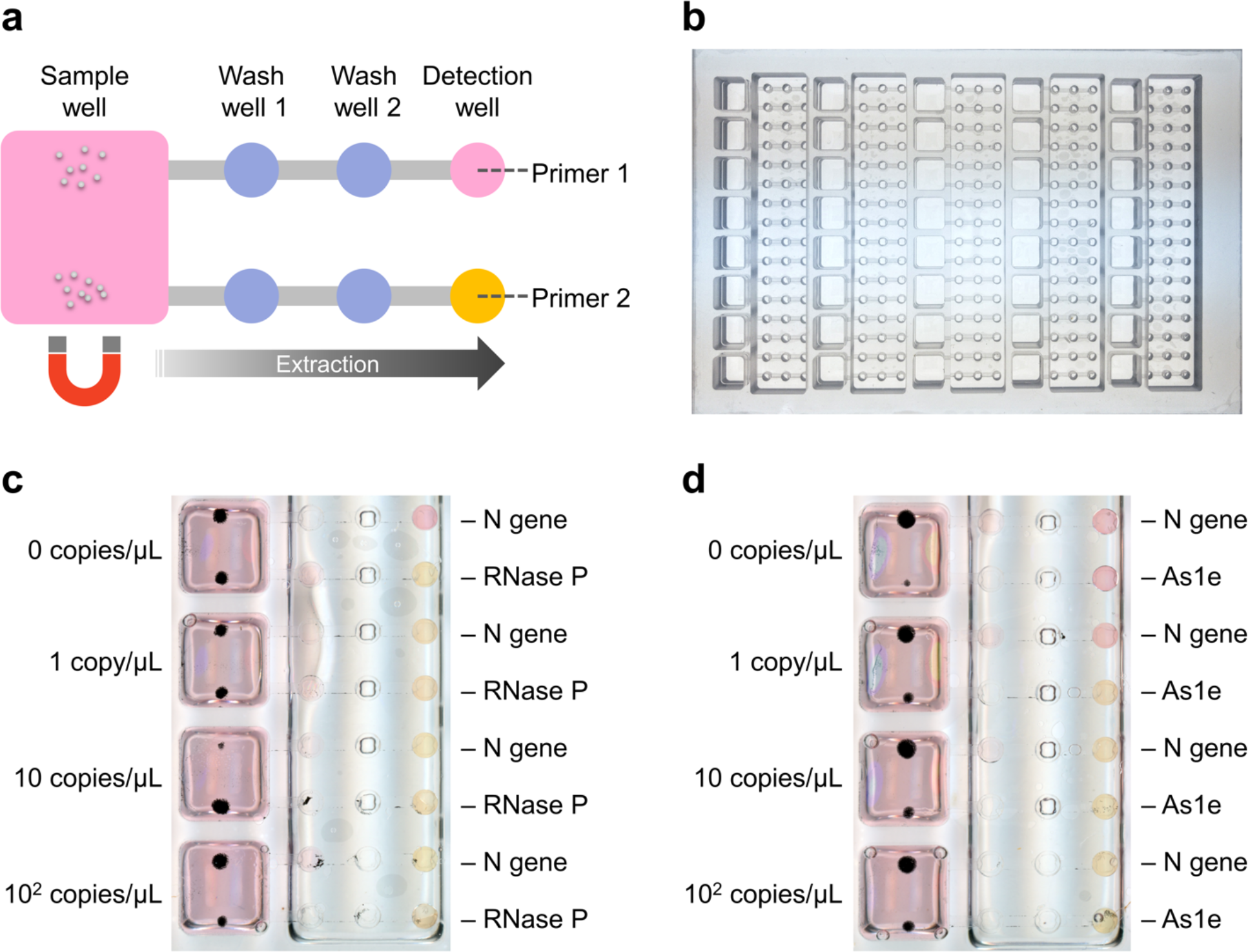
Dual-output OIL-TAS device enables simultaneous multiplexed detection from a single sample. **a** OIL-TAS can be designed with multiple extraction outputs (2 outputs shown here) leading from a single input sample well, enabling multiplexed detection of a sample by using different primers in the detection well. **b** Image of the dual-output OIL-TAS device which has the same footprint as a 96-well plate (127.76 mm x 85.48 mm). **c** Multiplexed detection of the SARS-CoV-2 N gene with an extraction positive control (RNase P). γ-irradiated inactivated SARS-CoV-2 particles were spiked into a cell suspension containing 10 A549 cells/μL in VTM to mimic a clinical sample. **d** Multiplexed detection of two different regions of the SARS-CoV-2 genome (As1e, which targets the open reading frame (Orf1a), and the N gene)

In summary, the OIL-TAS method provides a simple and fast integrated solution from sample to answer for SARS-CoV-2 testing. The OIL-TAS is advantageous in several ways: 1) Greatly reduced operation steps. RNA extraction and data acquisition is performed on the same device; 2) Reduction in biohazardous medical waste; 3) Extraction process is done under an oil overlay, preventing cross-contamination and aerosol formation; 4) Endpoint readout decouples reaction (isothermal amplification at 65 °C) from detection (flatbed scanner), thus freeing up instrument time for increased assay turnaround. A single operator can process a device (40 samples) within 30 minutes manually, which, including the 35 min incubation time adds up to a sample-to-result time of approx. 70 min; 5) Very low assay reagent and instrumentation cost; and 6) High specificity and good sensitivity. Importantly, although here we demonstrated the OIL-TAS using a manual operation protocol, the conventional plate format of the OIL-TAS device makes it very easily adaptable to an automated workflow using commercially available automated liquid handling systems and plate readers for further increased throughput. However, we acknowledge that even though OIL-TAS employs an established RNA extraction chemistry which likely makes it less sensitive to differences in sample type, further assay validation would be necessary to determine the real world performance and sensitivity of OIL-TAS in assaying clinical specimens across different sample types (i.e. anterior nares, mid-turbinate, nasopharyngeal, and oropharyngeal swabs, as well as saliva) and different sample storage/transport media.

## Methods

### Device Fabrication

Devices were milled out of 2.4 mm (for channels) and 4.5 mm (for top frame) thick polycarbonate sheets (LEXAN 9034, United States Plastic Corporation) using a computer numerical control (CNC) 3-axis mill (Tormach, PCNC 770). Following milling, the two pieces were thoroughly washed with 100% isopropyl alcohol and air dried using an air gun. The two milled pieces were then aligned and placed on top of a 0.005 inch (127 μm) thick polycarbonate sheet (TAP Plastics) and bonded together in a thermal press (Carver Press, 3889.1NE1001) with a pressure of 3000 Kg for 40 min. The bonded device was then treated with oxygen plasma for 2 min at 100 W (Diener Electronic Femto, Plasma Surface Technology). After plasma treatment, the device was placed in a vacuum desiccator with 2 trays (40 μL each) of PDMS silane (1,3’dichlorotetramethylsiloxane, Gelest, SID3372.0). The desiccator was then pumped down to vaporize and condense the PDMS silane onto the device surface at RT overnight to functionalize the device surface. The device was then thoroughly rinsed with 100% isopropyl alcohol then dried using an air gun to remove residual unattached PDMS silane.

### Reagent and sample preparation

Colorimetric LAMP master mix was prepared by mixing 100 μL of WarmStart Colorimetric LAMP 2X Master Mix (New England Biolabs), 20 μL of 10X LAMP primer mix, and 80 μL of nuclease free water (for 1 OIL-TAS device). LAMP primer sets were purchased from Integrated DNA Technologies, with primer sequences shown in Supplementary Table 1. The primers used include an N Gene primer^26^, As1e primer^29^, and RNase P primer^30^, The RNA lysis buffer was prepared using 4 M Guanidine thiocyanate, 10 mM MES (2-ethanesulfonic acid), 1% Triton X-100, with 1% ß-Mercaptoethanol added right before use. Qiagen MagAttract magnetic silica beads were diluted in 99% ethanol to reach an equivalent of 0.25 μL bead stock/extraction. For bead washing, 50% ethanol and nuclease free water was used for wash 1 and wash 2, respectively. Worth noting is that the rapid (∼1 s) and gentle nature of “washing” in the OIL-TAS method (dragging beads through a droplet) allowed us to use water as a washing solution while retaining good sensitivity. Gamma-irradiated inactivated SARS-CoV-2 viral particles (strain USA-WA1/2020) were obtained from the Biodefense and Emerging Infections Research Resources Repository (BEI), (cat# NR-52287) and serially diluted in VTM.

### Device operation (single output device)

To prepare the OIL-TAS device, 30 μL of silicone oil was added to each of the sample wells, which, due to the nature of the ELR surface treatment, the oil simultaneously wicks into the extraction channels and wells, forming an oil barrier to separate each well and also prevents sample evaporation. Using a p20 pipette, 4 μL of nuclease-free water, 50% ethanol, and LAMP master mix was sequentially added to washing well 2, washing well 1 and the detection well, respectively. SARS-CoV-2 sample lysis was performed on a 96-well round bottom plate. Briefly, 60 μL of RNA lysis buffer was added to each well, followed by addition of 30 μL sample. The plate was then placed on an orbital plate shaker for 5 min at 900 rpm to lyse the sample. After lysis, 60 μL of MagAttract beads/ethanol suspension (equivalent to 0.25 μL bead stock) was added to the wells and then placed on the plate shaker again for 5 min at 900 rpm for binding. (Note: the sample lysis and bead binding steps can also be performed in eppendorf tubes or other vessels). The bead/sample mixture with a total volume of 150 μL was transferred to the sample well of the OIL-TAS device to perform extraction. The OIL-TAS device was then placed onto a custom magnetic extractor plate containing an array of neodymium magnets with a diameter of 0.0625 inches (1.6 mm) (Rare-Earth Disc Magnets, MAGCRAFT) positioned under each well, which causes the beads to coalesce above each magnet. The OIL-TAS device was then slid from right to left over the magnetic extractor to perform RNA extraction, which transports each magnetic bead cluster from the sample well through the two washing wells and into the detection well, a process which takes less than 10 seconds. The beads were then left in the detection well for isothermal amplification by placing the device in an incubator/oven set at 65 °C for 35∼50 min. After amplification, beads were removed from the detection well (to allow for better colorimetric visualization) by placing the device on the magnetic extractor and sliding from left to right. Results of the colorimetric LAMP were then recorded by placing the device in a flatbed scanner (Epson Perfection V600 Photo). A positive reaction results in a color change of the droplet from pink to yellow, while a negative reaction droplet will remain pink.

### Device operation (dual output device)

Minor modifications to the operation protocol were made for the dual output OIL-TAS device (Fig. 5). The dual output OIL-TAS device was loaded with 50 μL of silicone oil to each of the sample wells. Using a p20 pipette, 4 μL of nuclease-free water, 50% ethanol, and LAMP master mix were sequentially added to washing well 2, washing well 1 and the detection well, respectively in the same manner as the single output device (except different primer sets were used in the two output detection wells). SARS-CoV-2 sample lysis and bead binding was performed in a 96-well deep well plate (or eppendorf tubes). 120 μL of RNA lysis buffer was added to each well of the well plate, followed by addition of 60 μL sample to the lysis buffer. The plate was then placed on an orbital plate shaker for 5 min at 900 rpm to lyse the sample. After lysis, 90 μL of MagAttract beads/ethanol suspension (equivalent to 0.5 μL bead stock) was added to the individual wells and then placed on the plate shaker again for 5 min at 900 rpm for binding. The bead/sample mixture with a total volume of 270 μL was transferred to the sample well of the OIL-TAS device to perform extraction. Magnetic extraction, amplification, and detection was performed as in the single output device.

### Quantification of carryover

Sample purification performance in the OIL-TAS process was evaluated by quantifying the amount of aqueous liquid carryover from one well to another following extraction. This was done by extracting magnetic beads through a fluorescent dye solution (acridine orange) as a model contaminant (Supplementary Fig. 2). In brief, a 4 μL droplet of 500 μg/mL acridine orange solution in water was added to washing well 1 (input droplet) of the OIL-TAS device, and a 4 μL droplet of deionized water was added to washing well 2 (output droplet). Carryover was defined by the amount of aqueous liquid that is carried over from the input droplet to the output droplet. In order to obtain a standard curve for fluorescence quantification, 4 μL droplets of 2-fold serially diluted acridine orange solution ranging from 10 μg/mL to 0 μg/mL were added to the detection wells. 100 μL of 1:400 diluted Qiagen MagAttract bead suspension in deionized water was added to the sample well, then extracted using the magnetic extractor through the acridine orange input droplets and into the water output droplets. The beads were subsequently removed from the output droplets by magnetic extraction in the opposite direction. The OIL-TAS device was then placed in a plate reader (PHERAstar FS, BMG Labtech), and the fluorescence intensity of the output droplets and standard curve droplets was measured with an excitation of 485 nm and emission of 520 nm. Amount of carryover was determined by fitting the fluorescence intensity of the output droplet to the serially diluted acridine orange standard curve.

## Data Availability

The data generated in this study are available within the article and its supplementary information.

## Acknowledgements

We would like to thank Dr. Brianna Mullins, Dr. Jay Warrick, and Dr. Scott Berry for helpful input and suggestions for the project. This study was funded by a UW WARF COVID-19 Accelerator Challenge award and the National Institutes of Health (NIH 1R01CA247479, NIH P30CA014520, NIH 1R01CA247479-01, NIH OD011106, NIH 5UL1TR002373-03).

## Contributions

D.S.J., T.D.J., D.M.D., and C.M.N. designed and performed experiments. D.J.B., D.H.O., and T.C.F. oversaw the study. D.S.J. and T.D.J. wrote the manuscript with input from all authors.

## Corresponding author

Correspondence to David J. Beebe.

