## Supplementary Information for "Oil Immersed Lossless Total Analysis System (OIL-TAS): Integrated RNA Extraction and Detection for SARS-CoV-2 Testing"

### Table of contents:

**Supplementary Fig. 1:** Graphical operation protocol for SARS-CoV-2 testing using OIL-TAS....... S-2

**Supplementary Fig. 2:** Extraction carryover of OIL-TAS ........ S-2

**Supplementary Fig. 3:** Droplet stability of OIL-TAS ........ S-3

**Supplementary Fig. 4:** Performance of OIL-TAS using a reagent freshly loaded device compared to a reagent pre-loaded device that was frozen for a week at -20 ºC ........ S-3

**Supplementary Table 1:** LAMP primer sequences ........ S-4

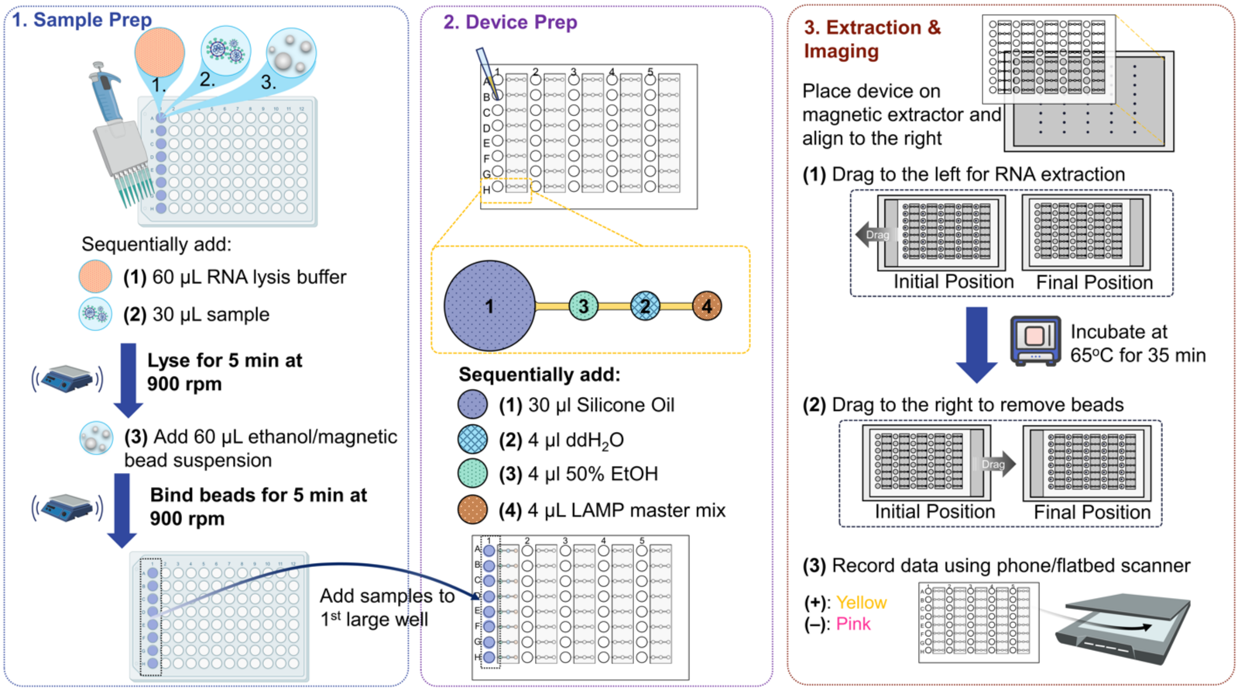


**Supplementary Fig. 1** Graphical operation protocol for SARS-CoV-2 testing using OIL-TAS.

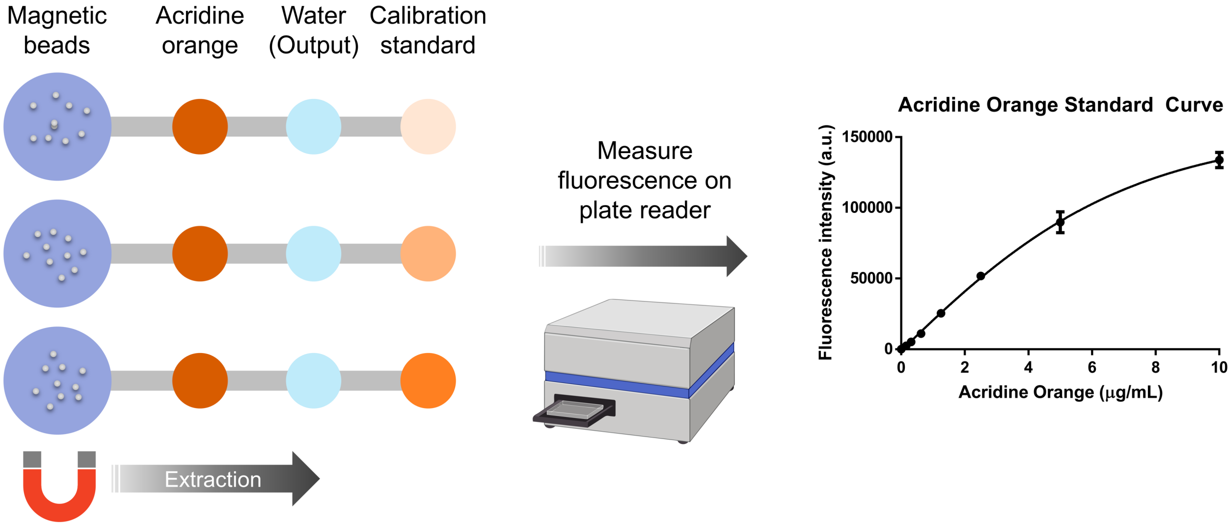


**Supplementary Fig. 2** Extraction carryover of OIL-TAS. The amount of carryover was calculated by fitting the fluorescence intensity of the water (output) droplet to the serially diluted acridine orange standard curve.

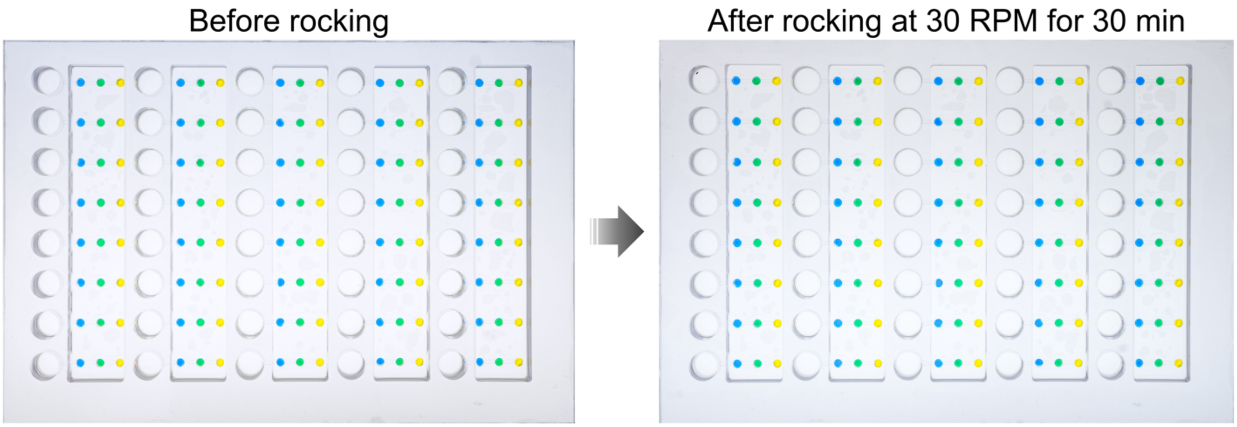


**Supplementary Fig. 3** Droplet stability of OIL-TAS. No droplets were dislodged from the wells in the device after rocking on a rocking platform shaker at 30 RPM for 30 min.

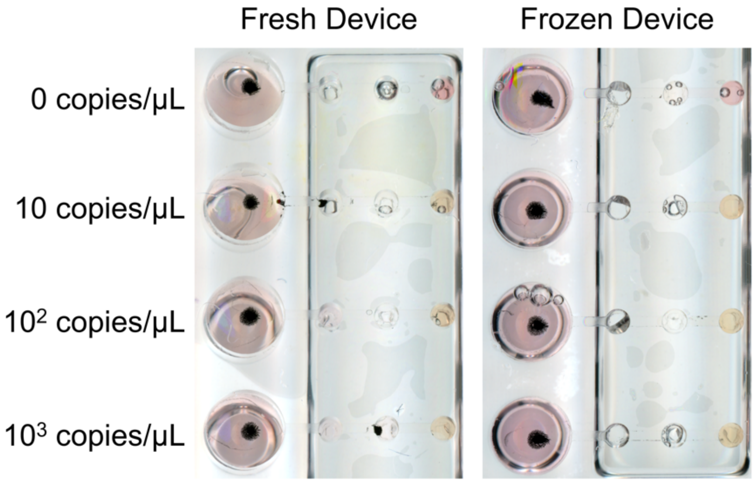


**Supplementary Fig. 4** Performance of OIL-TAS using a reagent freshly loaded device (left) compared to a reagent pre-loaded device that was frozen for a week at -20 ºC (right).

**Supplementary Table 1:** LAMP primer sequences

| Primer Set | Sequence | Reference |
|---|---|---|
| **N gene** | | 26 |
| F3 | AACACAAGCTTTCGGCAG | |
| B3 | GAAATTTGGATCTTTGTCATCC | |
| FIP | TGCGGCCAATGTTTGTAATCAGCCAAGGAAATTTTGGGGAC | |
| BIP | CGCATTGGCATGGAAGTCACTTTGATGGCACCTGTGTAG | |
| LF | TTCCTTGTCTGATTAGTTC | |
| LB | ACCTTCGGGAACGTGGTT | |
| **As1e** | | 29 |
| F3 | CGGTGGACAAATTGTCAC | |
| B3 | CTTCTCTGGATTTAACACACTT | |
| FIP | TCAGCACACAAAGCCAAAAATTTATTTTTCTGTGCAAAGGAAATTAAGGAG | |
| BIP | TATTGGTGGAGCTAAACTTAAAGCCTTTTCTGTACAATCCCTTTGAGTG | |
| LF | TTACAAGCTTAAAGAATGTCTGAACACT | |
| LB | TTGAATTTAGGTGAAACATTTGTCACG | |
| **RNase P** | | 30 |
| F3 | TTGATGAGCTGGAGCCA | |
| B3 | CACCCTCAATGCAGAGTC | |
| FIP | GTGTGACCCTGAAGACTCGGTTTTAGCCACTGACTCGGATC | |
| BIP | CCTCCGTGATATGGCTCTTCGTTTTTTTCTTACATGGCTCTGGTC | |
| LF | ATGTGGATGGCTGAGTTGTT | |
| LB | CATGCTGAGTACTGGACCTC | |
